# Previously infected vaccinees broadly neutralize SARS-CoV-2 variants

**DOI:** 10.1101/2021.04.25.21256049

**Authors:** Hans C. Leier, Timothy A. Bates, Zoe L. Lyski, Savannah K. McBride, David X. Lee, Felicity J. Coulter, James R. Goodman, Zhengchun Lu, Marcel E. Curlin, William B. Messer, Fikadu G. Tafesse

**Affiliations:** Department of Molecular Microbiology & Immunology, Oregon Health & Science University; Portland, OR 97239, United States; Medical Scientist Training Program, Oregon Health & Science University; Portland, OR 97239, United States; Division of Infectious Diseases, Oregon Health & Science University; Portland, OR 97239, United States

## Abstract

We compared the serum neutralizing antibody titers before and after two doses of the BNT162b2 COVID-19 vaccine in ten individuals who recovered from SARS-CoV-2 infection prior to vaccination to 20 individuals with no history of infection, against clinical isolates of B.1.1.7, B.1.351, P.1, and the original SARS-CoV-2 virus. Vaccination boosted pre-existing levels of anti-SARS-CoV-2 spike antibodies 10-fold in previously infected individuals, but not to levels significantly higher than those of uninfected vaccinees. However, neutralizing antibody titers increased in previously infected vaccinees relative to uninfected vaccinees against every variant tested: 5.2-fold against B.1.1.7, 6.5-fold against B.1.351, 4.3-fold against P.1, and 3.4-fold against original SARS-CoV-2. Our study indicates that a first-generation COVID-19 vaccine provides broad protection from SARS-CoV-2variants in individuals with previous infection.

## Main

In recent months, multiple coronavirus disease 2019 (COVID-19) vaccine candidates have successfully concluded phase 3 trials^1^, with the three candidates authorized for emergency use by the U.S. Food and Drug Administration reporting efficacies of 95% (BNT162b2 [Pfizer-BioNTech]), 94% (mRNA-1273 [Moderna]), and 66% (Ad26.COV2.S [Janssen])^2-4^. When combined with the substantial portion of many communities estimated to have gained natural immunity through infection with severe acute respiratory syndrome coronavirus 2 (SARS-CoV-2)^5^, the rollout of safe and effective vaccines has raised the possibility that high levels of population immunity could soon be reached. Clouding this prospect is the emergence and global spread of SARS-CoV-2 variants of concern (VOCs), such as those first identified in the United Kingdom (lineage B.1.1.7)^6^, South Africa (B.1.351)^7^, Brazil (P.1)^8^, and California (B.1.429)^9^. Most VOCs possess partially overlapping combinations of spike mutations that enhance binding to the SARS-CoV-2 cellular receptor angiotensin-converting enzyme 2 (ACE2), increasing transmissibility^10^ (Supplementary Table 1). More concerning has been the emergence of spike mutations with the potential to escape neutralizing antibodies raised against earlier lineages of SARS-CoV-2 through infection with an original linage or by first-generation COVID-19 vaccines^11-13^. Recent population studies have validated these findings, showing surges of reinfections in regions with extensive transmission of B.1.35^17,14^ and P.1^8^, and large declines in vaccine efficacy against B.1.351^14-16^.

To investigate whether vaccination of individuals previously infected by SARS-CoV-2 confers greater protection from VOCs than vaccination of individuals with no evidence of previous infection, in a cohort of BNT162b2 vaccinees (Supplementary Table 2) we identified a group of 10 study participants who had received a positive COVID-19 PCR test result prior to vaccination, along with an age- and sex-balanced group of 20 participants who had not. While vaccination of previously infected individuals boosted the 50% maximal effective concentration (EC50) of antibodies against the immunodominant SARS-CoV-2 spike receptor-binding domain (RBD) ten-fold (pre-vaccination geometric mean titer [GMT], 82.15; post-vaccination GMT, 823.3), the vaccine-elicited antibody titers of uninfected individuals (GMT, 699.5) were not significantly lower (Fig. 1A). Similarly, post-vaccination levels of RBD-binding IgG (Fig. 1B) and IgA (Fig. 1C) did not differ significantly between the two groups.

**Figure 1.**
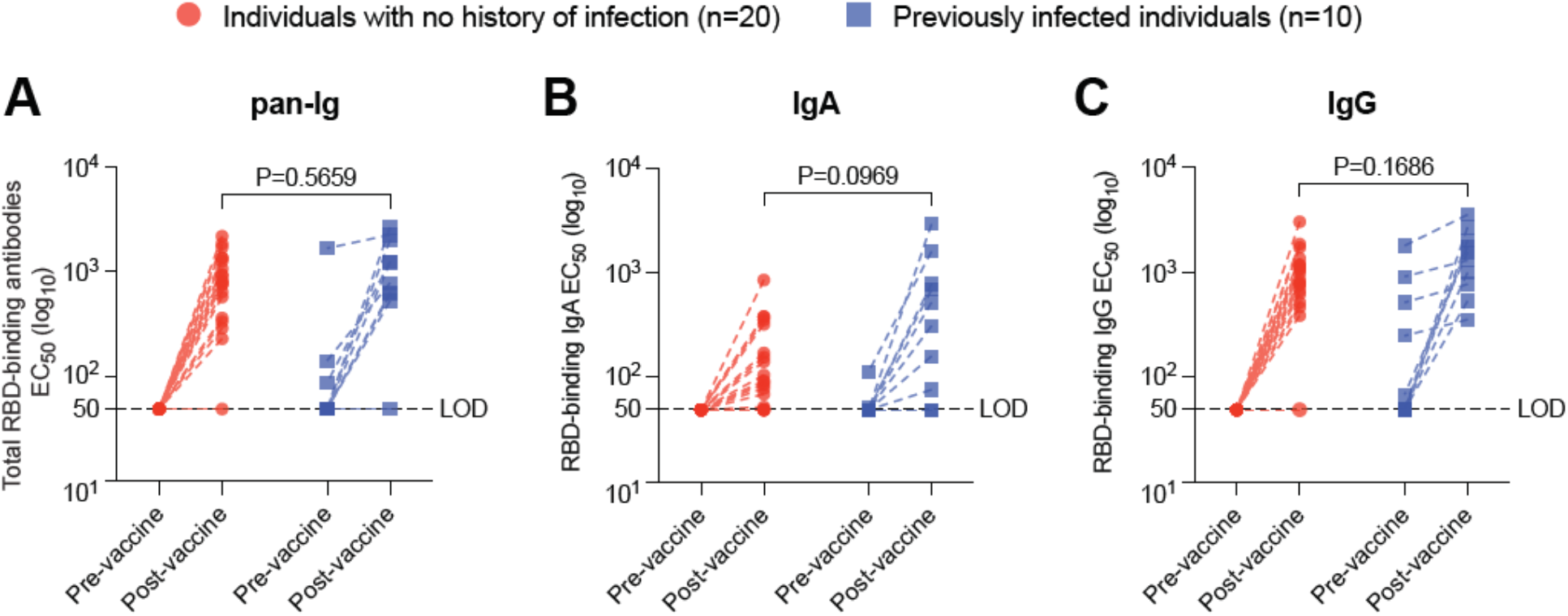
Anti-SARS-CoV-2 antibody levels elicited by vaccination and natural infection. (**A**) Half maximal effective concentration (EC50) of total pan-Ig antibodies specific to the spike RBD were measured by enzyme-linked immunosorbent assay (ELISA) in serum collected from donors previously infected with SARS-CoV-2 pre- and post-vaccination with the BNT162b2 vaccine. (**B**) EC50 of RBD-binding IgA. (**C**) EC50 of RBD-binding IgG. Statistical comparisons were made using the Wilcoxon rank-sum test. LOD denotes limit of detection.

We then measured the pre- and post-vaccination neutralizing activity of the two groups of sera against an early SARS-CoV-2 isolate (USA-WA1/2020) and isolates of B.1.1.7, B.1.351, and P.1 (Fig. 2). Pre-vaccination sera from previously infected participants provided higher levels of neutralization against USA-WA1/2020 (GMT, 39.0) than against the three VOCs (GMT, 25.7 for B.1.1.7; GMT<20 for B.1.351; GMT, 31.2 for P.1), consistent with previous reports of convalescent sera^12,17^. Similarly, post-vaccination sera from uninfected participants showed greater neutralization of USA-WA1/2020 than of the VOCs (GMT, 578.6 for USA-WA1/2020; 223.0 for B.1.1.7; 47.5 for B.1.351; 171.9 for P.1). However, post-vaccination serum from previously infected individuals possessed significantly higher neutralizing activity against every SARS-CoV-2 lineage relative to post-vaccination serum from uninfected participants: neutralizing antibody titers increased by a factor of 3.5 against USA-WA1/2020 (95% confidence interval [CI], 2.8 to 4.0); by a factor of 5.2 against B.1.1.7 (95% CI, 2.37 to 9.8); by a factor of 6.5 against B.1.351 (95% CI, 3.4 to 12.3); and by a factor of 4.3 against P.1 (95% CI, 2.8 to 6.5). Notably, there was no significant difference (P=0.2736, Wilcoxon rank-sum test) between the post-vaccination neutralizing antibody titers of previously infected participants against B.1.351 (GMT, 307.3; 95% CI, 91.0 to 1038) and those of uninfected participants against USA-WA1/2020 (GMT, 578.6; GMT, 332.5 to 1007), suggesting that first-generation COVID-19 vaccines could retain near-complete efficacy against even the most resistant VOCs when administered following natural infection.

**Figure 2.**
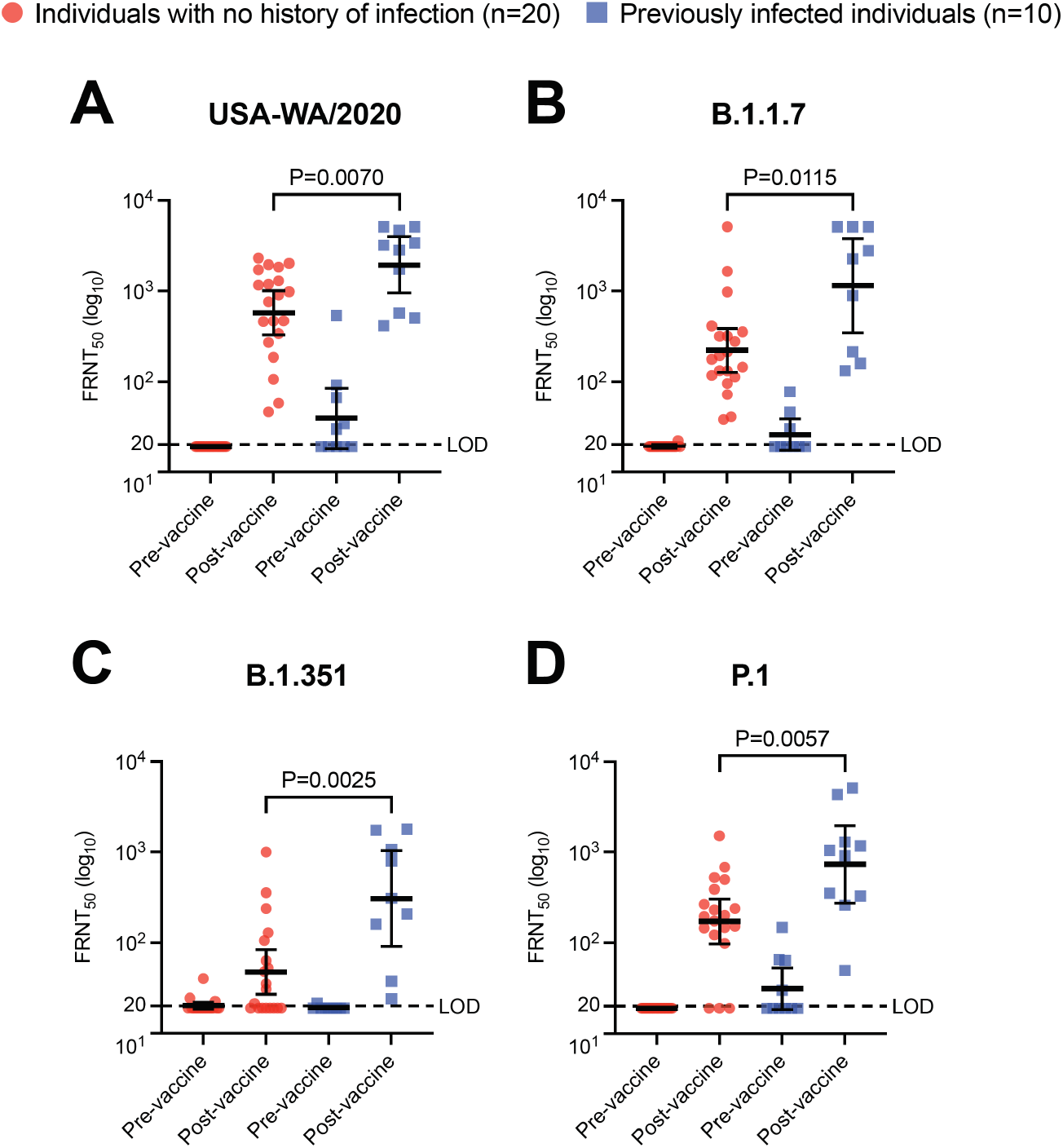
Immunity from natural SARS-CoV-2 infection boosted by subsequent vaccination broadens protection against variants of concern. Neutralization of SARS-CoV-2 variants by pre-and post-vaccination sera collected from previously infected and naïve individuals. Shown are the results of a 50% focus reduction neutralization testing (FRNT50) for an early SARS-CoV-2 isolate USA-WA1/2020 (**A**) and VOC clinical isolates of B.1.1.7 (**B**), B.1.351 (**C**), and P.1 (**D**). Horizontal bars represent geometric mean titer and I bars represent 95% confidence intervals. Statistical comparisons were made using the Wilcoxon rank-sum test. LOD denotes limit of detection.

Overall, our findings provide important evidence for broad and potent neutralizing antibody responses against emerging SARS-CoV-2 variants, even with exposure to only wildtype SARS-CoV-2 antigen. This reinforces a recent report that natural infection with B.1.351 elicits a similar cross-reactive neutralizing antibody response against B.1.351, P.1, and original SARS-CoV-2^18^. While these and other laboratory results must be validated by ongoing population-level studies, they indicate a novel role for COVID-19 vaccines in protecting hard-hit populations from future waves of the pandemic.

## Data Availability

All data in this manuscript is available upon request from the corresponding authors, M.E.C., W.B.M., and F.G.T.

## Acknowledgments

The authors thank the clinical staff at Oregon Health & Science University who assisted in enrolling study participants and collecting the serum samples used in this report.

## Data and materials availability

All data are available in the main text or the supplementary materials.

## Supplementary Materials for

### Materials and Methods

#### Cohort

This study was conducted in accordance with the Oregon Health & Science University (OHSU) Institutional Review Board (IRB#00022511). Study participants were enrolled at OHSU immediately after receiving the first dose of the BNT162b2 (Pfizer-BioNTech) COVID-19 vaccine. After receiving informed consent, 4-6 mL whole blood was drawn and centrifuged 10 minutes at 1000 x g. A second blood sample was obtained at least 14 days after participants received their second vaccine dose. Serum was stored at - 20°C until heat inactivation in preparation for FRNTs and ELISAs. n=10 participants were identified with a positive COVID-19 PCR test prior to vaccination and n=20 age- and sex-balanced participants who did not have a history of positive PCR test or RBD-binding antibodies in an ELISA. See Supplementary Table 1 for more details of the cohort.

#### ELISA

The 96-well ELISA plates (Corning, Cat# 3590) were coated with recombinant RBD protein prepared as previously described^19^ at a concentration of 1 ug/mL in PBS and incubated overnight at 4°C. Coating antigen was removed, plates were washed once with PBST containing 0.05% Tween (wash buffer) and blocked for 1 hour at RT with 5% milk prepared in PBST containing 0.05% Tween (dilution buffer). 100 mL of 1:20 (pre-vaccine), or 1:200 (post-vaccine) dilution of serum in dilution buffer was added to each starting well. Three-fold serial dilutions were performed in dilution buffer. Plates were incubated at room temperature for 1 hour. The plates were washed 3 times with wash buffer and 100 mL of 1:10,000 dilution of anti-human GOXHU IGG/A/M HRP (Invitrogen, Cat# A18847) detection antibody was added and incubated at room temperature for 1 hour. After washing the plates 3 times with wash buffer, 100 mL of colorimetric detection reagent containing 0.4 mg/ml o-phenylenediamine and 0.01% hydrogen 45 peroxide in 0.05 M citrate buffer (pH 5) were added and the reaction was stopped after 20 minutes by the addition of 100 mL 1 M HCl. Optical density (OD) at 492 nm was measured using a CLARIOstar plate reader. Plates were normalized, background removed, and endpoint antibody titers determined by the highest dilution with a positive signal, defined as 4-fold above background. Final endpoint titer values below the detection limit of 20 were set to 19.

#### Viruses

SARS-CoV-2 isolates were obtained from BEI Resources (Supplementary Table 2): Isolate USA/CA_CDC_5574/2020 [B.1.1.7] (BEI Resources NR-54011); Isolate hCoV-54 19/South Africa/KRISP-K005325/2020 [B.1.351] (BEI Resources NR-54009); Isolate hCoV-19/Japan/TY7-503/2021 [P.1] (BEI Resources NR-54982); and Isolate USA-WA1/2020 [early isolate] (BEI Resources NR-52281). Isolates were propagated and titrated in Vero E6 cells as previously described^20^.

#### FRNT

Serial dilutions of patient sera and virus neutralization were carried out in biological duplicate in a 96-well plate format. Briefly, each sample was added in duplicate 1:10 to dilution media (Opti-MEM, 2% FBS), and four-fold serial dilutions were made for a range of 1:10 – 1:2560. An equal volume of dilution media containing 100 FFU of SARS-CoV-2 was added to each well (final dilutions of sera, 1:20–1:5120) and incubated 1 h at 37°C. The virus-serum mixtures were then added to monolayers of Vero E6 cells, incubated with occasional rocking 1 h at 37°C, and covered with overlay media (Opti-MEM, 2% FBS, 1% methylcellulose). Overlay media was then removed, and plates were fixed for 1 h in 4% paraformaldehyde in PBS. Foci were developed as described. Plates were imaged with a CTL Immunospot Analyzer, then foci were counted using CTL ImmunoSpot (7.0.26.0) Percent neutralization values for FRNT50 were compiled and analyzed using python (v3.7.6) with numpy (v1.18.1), scipy (v1.4.1), and pandas (v1.0.1) data analysis libraries. Replicate data were fit with a three-parameter logistic model and final FRNT50 values below the lowest dilution of 20 were set to 19 while values above the maximum dilution of 5120 were set to 5121. FRNT50 curves were plotted using python with the Matplotlib (v3.1.3) data visualization library.

### Statistical analysis

Aggregated ELISA endpoint titers and FRNT50 values were analyzed in GraphPad Prism (v9.0.2). Comparisons were performed using the Wilcoxon rank-sum test. Reported group averages are geometric means, with error bars representing 95% confidence intervals.

**Supplementary Table 1.** List of mutations in B.1.1.7, B.1.351 and P.1 SARS-CoV-89 2 clinical isolates.*

| Lineage | GISAID Clade | GISAID ID | Spike mutations (RBD highlighted) | Non-Spike mutations |
| --- | --- | --- | --- | --- |
| **B.1.1.7** | GR | EPI_ISL_683466 | H69del, V70del, Y145del, **N501Y**, A570D, D614G, P681H, T716I, S982A, D1118H | N D3L, N G204R, N R203K, N S235F, NS8 Q27stop, NS8 R52I, NS8 Y73C, NSP3 A890D, NSP3 A1305V, NSP3 I1412T, NSP3 T183I, NSP6 F108del, NSP6 G107del, NSP6 S106del, NSP12 P323L, NSP13 K460R, NSP14 E347G |
| **B.1.351** | GH | EPI_ISL_678570 | D80A, D215G, L242del, A243del, L244del, **K417N**, **E484K**, **N501Y**, D614G, A701V | E P71L, N T205I, NS3 Q57H, NS3 S171L, NSP2 T85I, NSP3 K837N, NSP5 K90R, NSP6 F108del, NSP6 G107del, NSP6 S106del, NSP12 P323L |
| **P.1** |  |  | D138Y, D614G, **K417T**, **E484K**, **N501Y**, G181V, H655Y, L18F, P26S, R190S, T20N, T1027I, V1176F | N G204R, N P80R, N R203K, NS3 S253P, NS8 E92K, NSP3 K977Q, NSP3 S370L, NSP3 T186A, NSP6 F108del, NSP6 G107del, NSP6 S106del, NSP12 P323L, NSP13 E341D |

\*Obtained from BEI Resources

**Supplementary Table 2.** Description of cohort.

|  | **Previously infected**<br>**(n=10)** | **No history of infection**<br>**(n=20)** |
| --- | --- | --- |
| **Sex – no. (%)** |  |  |
| Female | 6 (60) | 15 (75) |
| Male | 4 (40) | 5 (25) |
| **Median age – yrs (range)** | 36.5 (23-61) | 44 (23-60) |
| **Median time between vaccine**<br>**doses – days (range)** | 22 (20-25) | 21 (21-32) |
| **Median time from second**<br>**vaccine dose to second**<br>**serum sample – days (range)** | 18 (14-28) | 17 (14-28) |
| **Median time from positive**<br>**COVID-19 PCR test to first**<br>**vaccine dose – days (range)** | 52 (13-302) | NA |

